# Clinical Validation of AI/ML Based Model for Down Syndrome Detection Through Graphical Analysis of Facial Dysmorphic Features

**DOI:** 10.1101/2025.10.26.25338853

**Authors:** Binoy V Shah, Saanvi Mehra, Justin Guerra, Ankur Sethi, Ratna Dua Puri, Somashekhar M Nimbalkar

**Author notes:** Contributions: BVS: Data Curation, Formal Analysis, Writing – Original Draft Preparation, Writing – Review & Editing, Validation, and Visualization. SM: Data Curation, Formal Analysis, Writing – Original Draft Preparation, Writing – Review & Editing, Validation, and Visualization. JG: Writing – Original Draft Preparation, Writing – Review & Editing AS: Conceptualization, Funding Acquisition, Investigation, Methodology, Project Administration, Resources, Supervision, Validation, and Visualization. RDP: Project Administration, Resources, Supervision, Validation, and Visualization. SMN: Conceptualization, Funding Acquisition, Investigation, Methodology, Project Administration, Resources, Supervision, Validation, and Visualization.

## Abstract

**Background and objective:** Down Syndrome is associated with high mortality in India, due to non-diagnosis/ late-diagnosis resource constraints. The objective of the present study was to test Clinical Validity of the Google Cloud AutoML Vision Image Classification Down Syndrome Detection model in real-life situations – i.e., does this model work in the hands of laymen (parents, guardians, end-user) with the same accuracy (98%+ in a controlled experiment).

**Methods:** This multi-label cross-sectional study was conducted in the Neonatal and Paediatric unit of a tertiary care hospital between August 2021 and September 2022. The participants consisted of 104 children aged 5 days - 18 years of two categories - those exhibiting facial dysmorphic features indicative of Down Syndrome on visual inspection, and those not exhibiting said features. Images were collected after written informed consent using the Down Syndrome Detection application. The outcomes recorded were the efficiency of the model in detection of Down Syndrome based of the images collected.

**Results:** The CloudML model trained with 104 images initially achieved: Sensitivity - 100%, specificity - 80%, Average Precision - 96.6%, precision - 86.67%, and Recall - 92.86% (Precision and Recall are calculated at a confidence threshold of 0.5) This Indo-specific Machine Learning model, specifically trained and tested on Indian children, shows remarkable accuracy in the diagnosis of Indian Down Syndrome positive neonates. On adjustment of software parameters (the confidence threshold of prediction), the technique can deliver highly accurate Down Syndrome diagnosis with a 100% Sensitivity, at the expense of false positives that may be ruled out through further confirmatory testing.

**Conclusions:** This diagnostic algorithm is a reliable preliminary postnatal screening tool and can be deployed in resource-limited settings where genetic testing is neither affordable nor readily accessible. False positives can be ruled out through subsequent testing.

## INTRODUCTION

Down Syndrome is a frequently occurring congenital anomaly worldwide, with an estimated prevalence ranging from 1 in 800 to 1 in 1200 live births. The pooled prevalence of congenital anomalies in India was found to be 184.48 per 10,000 births, but there is a lack of nationwide data specifically for Down syndrome, which might be due to presence of economically and anthropologically diverse population (1). Children with Down Syndrome are at increased risk of Congenital Heart Defects, developing Pulmonary Hypertension, and other medical conditions related to hearing and vision; many of these conditions are manageable if detected early (2). Despite of the Government’s best efforts, the genetic disease is associated with high mortality in India (3). Artificial Intelligence has made significant strides over the last decade and is now being extensively used for a variety of purposes, especially for image and pattern recognition across a variety of domains (4-6). The present study employs Machine Learning to facilitate the recognition of DS-related facial dysmorphic features in neonates of Indian origin.

The prenatal assessment of DS pregnancy risk is based on ultrasonic markers of foetal growth and biological markers present in maternal serum. The Detection Rate and False Positive Rate for these methods continues to pose a challenge to early detection. One of the major challenges in India is access to healthcare and affordability. The healthcare system heavily relies on the private sector, which has resulted in the concentration of medical infrastructure in large metropolitan areas. However, the majority of the population resides in rural areas, leading to limited access to inpatient and outpatient facilities for most people living in these areas (8-10). The combination of poverty, limited awareness, and low education levels exacerbates the issue, as many deliveries still take place at home without the presence of qualified healthcare professionals or access to emergency medical facilities (11, 12). Quality of care being provided in some parts of the country is also an important factor in newborn survival (13). The stress caused by the waiting period between taking a test and receiving the results could be a factor that leads some high-risk couples to choose not to be tested (14).

The primary markers of a Down Syndrome (DS) baby can be flagged before birth through an ultrasound scan in the first trimester between 11 to 14 weeks, which measures nuchal translucency, which continues to be a luxury for more than 90% of Indian parents. Even for those with access to medical facilities, an ultrasound scan is rarely conclusive with a detection rate of 70% or less and only indicates an increased risk of DS (15). An ultrasound is to be followed up by maternal serum tests and other expensive tests.

Maternal serum screening in the second trimester (between 14 and 20 weeks) is another method for risk assessment of DS. This includes a double test [maternal serum alpha-fetoprotein (MSAFP) + beta subunit of human chorionic gonadotropin (β-hCG)], a triple test [double test + unconjugated estriol (uE3)], and a quadruple test (triple test + inhibin A). The Detection Rate and False Positive rate of these tests have not been very promising for risk assessment. The detection rate (DR) for the Double test is 57, for the triple tests, the DR is ∼69% and the quadruple test achieves a DR of up to 81% (15). Tests such as Amniocentesis and Fetal Heart Check-ups are expensive. Besides cost, Amniocentesis is associated with a small probability of miscarriage as well as foetal deformation - leaving parents-to-be with some uncomfortable and difficult choices (16-18). Karyotyping and Fluorescence in situ hybridization (FISH) [19] tests are reliable postnatal tests that the Indian Government is subsidising and promoting vigorously. However, carrying out this test in every remote, inaccessible corner of the country continues to be a challenge. Parents who do not utilize healthcare services for childbirth are unlikely to seek out invasive and costly tests through the same system.

The challenges with current prenatal testing protocol include lack of awareness in remote areas, prohibitively expensive cost of testing, especially for the underprivileged, a lack of access to medical infrastructure/ ultrasound facilities in rural areas, inconclusive results from primary prenatal testing through ultrasound, misuse of ultrasound testing for prenatal gender determination and gender selection, poor DR and FPR of the blood serum markers, risk to the foetus with follow-on prenatal testing such as Amniocentesis, and a long waiting time for results with follow-on tests. The objective of this research was to Clinically Validate the Machine Learning model (7) for first-level screening of Down Syndrome among children of Indian origin and to assess the suitability of the Google Cloud AutoML Vision Image Classification Model as a preliminary diagnostic aid.

## METHODS

This multi-label cross-sectional study was conducted in the Neonatal and Paediatric unit of a tertiary care hospital between August 2021 and September 2022. The research was approved by the institutional ethics committee. Written informed consent was obtained from parents of both Down Syndrome positive and negative patients prior to image and data collection.

A four-step methodological approach was adopted to develop and validate the Down Syndrome diagnostic algorithm/ aid for paediatric and neonatal patients suspected to be afflicted with the genetic disorder (**figure 1**). For validation, we calculated the predictive sensitivity and specificity on the basis of the precision, recall, and accuracy results returned by the model (20).

**Figure 1:**
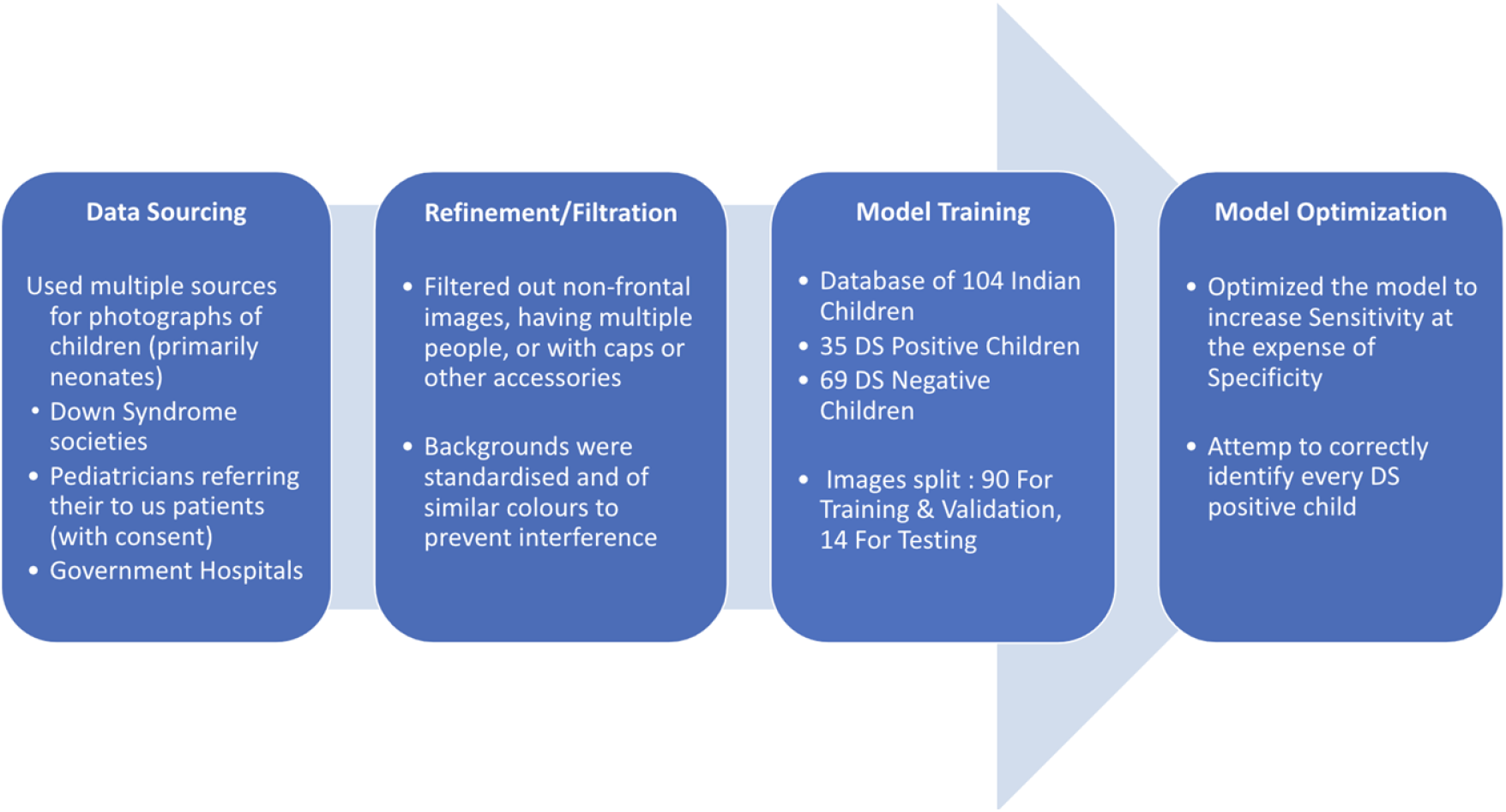
Flow diagram of Four-step process for AI/ML based Down Syndrome Detection.

A list of signs and symptoms for Down Syndrome was created through a literature review, which could allow paediatricians, neonatologists, and geneticists to make a preliminary diagnosis based on visual inspection alone, without needing to conduct genetic testing. Images were collected of all infants and children, even those afflicted with other diseases, as long as their facial features were not affected by the disease. For patients coming in specifically for Down Syndrome diagnosis and treatment, facial photographs were collected, and the diagnosis was later confirmed (or contradicted) by their respective Karyotyping test reports.

All collected images were standardised to filter out non-frontal images, images having multiple people, or images with caps or other accessories in order to ensure that these factors do not skew the predictions made by the Machine Learning model, adversely affecting the diagnostic capabilities of the Artificial Intelligence algorithm by impacting the very grounds on which it makes the predictions in question. It is important to note that all images were clicked with plain, most frequently white backgrounds to ensure that no external factor influences the decision-making of the model’s algorithm.

The final database amounted to 104 images, with a training and testing split. 90 images were used by the model for Training and Validation. The remaining 14 images were used for testing purposes. Of particular relevance was the group of patients, which consisted of infants who were suspected of having Down Syndrome but were confirmed not to have the genetic disorder on further genetic testing. This group was closely monitored to identify common characteristics that led to an incorrect initial diagnosis.

All diagnostic test results were later tabulated to calculate the total numbers of Down Syndrome positive and Down Syndrome negative subjects. The model was then trained on the 90 images of both Down Syndrome positive and Down Syndrome Negative children, with it being a 35:69 split. This split has not been calculated to yield a specific result or accuracy - it is simply the number of suitable images and data-points collected for the purpose of this study. We recorded all Down Syndrome positive patients that arrived at the care centre, but only a few suitable neonatal candidates emerged when it came to the collection of Down Syndrome negative subjects.

Optimising the model to prioritise Sensitivity over Specificity was pertinent and in alignment with the objective of the study. It was our purpose to ensure (to the greatest extent possible) that every Down Syndrome positive case was correctly identified, even if a few false positives had to be brought in for contradiction of the preliminary diagnosis.

### Statistical Analysis

The data was analysed using Microsoft Excel (2019) and Google Cloud AutoML’s embedded graphing and confusion matrix-building tools. The facial dysmorphic properties were analysed and given appropriate weightage internally by the model itself, without any manual intervention. The model assigns a score between 0 and 1 (both limits included) to determine the likeliness of the diagnostic result, with 1 being completely definitive and 0 being completely contradictory. Predictive Validity, Predictive Sensitivity, and Predictive Specificity was calculated based on the Precision-Recall trade-off graphs. Average precision is used as a measure of accuracy of the model (21). Dashboard precision and recall values were calculated at a confidence threshold of 0.5. Average precision was calculated by determining the area under the Precision-Recall trade-off curve.

### Ethical Approval

The study was approved by Ethics Committee, Institutional Ethics Committee-2 Bhaikaka University, Karamsad, Anand, Gujarat on 10^th^ March 2022 (Clearance number IEC/ BU/2022/Ex. 01). The participants’ mothers and other family members were explained the procedure of the study, their roles, and the outcome. Written informed consent (paper form) was obtained from each participant’s mother.

## RESULTS

With the initial confidence threshold (22) at 0.5, the model yielded the following Confusion Matrix, and achieved a Precision of 86.67%, Recall of 92.86%, sensitivity of 100%, specificity of 80%, and Average Precision of 96.6% **(table 1)**. The graph **(figure 2)** highlights that sensitivity has already been maximised at the standard confidence threshold of 0.5-eliminating all false negatives. Whilst there are still false positives, they can be ruled out through subsequent testing. However, 100% of the DS children can be correctly identified (equivalent to Sensitivity of Down Syndrome test),

**Table 1.**
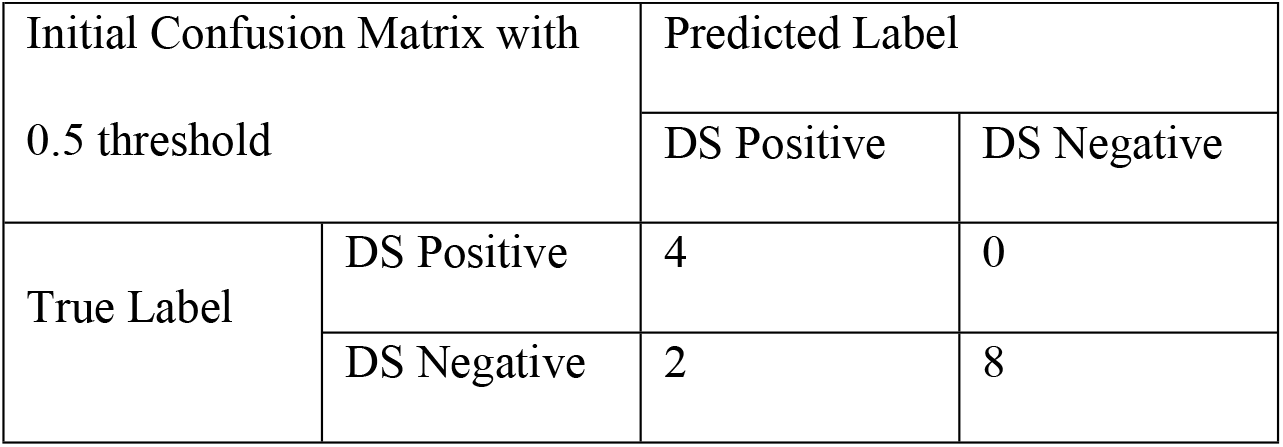

**Figure 3: Precision v/s Recall Trade-off for 0.05 confidence threshold**
| Table 1 |  |  |  |
| --- | --- | --- | --- |
| Initial Confusion Matrix with<br>0.5 threshold |  | Predicted Label |  |
|  |  | DS Positive | DS Negative |
| True Label | DS Positive | 4 | 0 |
|  | DS Negative | 2 | 8 |

**Figure 2:**
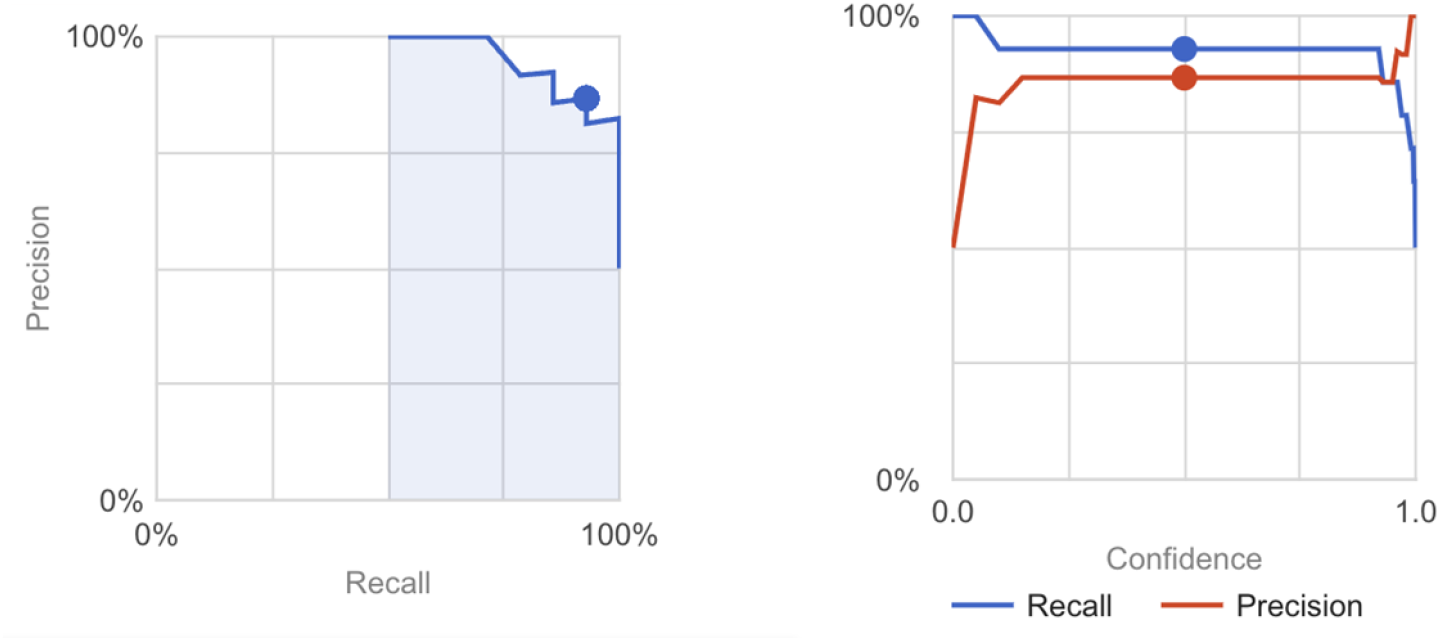
Precision v/s Recall Trade-off for 0.5 confidence threshold.

As can be seen from the graph, for precision v/s recall tradeoff for various confidence threshold levels **(figure 3)**, upon changing the threshold to 0.05 (moving from the red dot to the blue dot below), the Recall (Sensitivity) value could be improved to almost perfect levels without too much compromise on precision. Changing the threshold yielded the following Confusion Matrix **(table 2)**. At a confidence threshold level of 0.05, we reach precision of 82.35%, Recall of 100%, sensitivity of 100%, specificity of 80%, and Average Precision of 85.71%.

**Table 2.**

| Table 2 |  |  |  |
| --- | --- | --- | --- |
| Initial Confusion Matrix with<br>0.05 threshold |  | Predicted Label |  |
|  |  | DS Positive | DS Negative |
| True Label | DS Positive | 4 | 0 |
|  | DS Negative | 2 | 8 |

**Figure 3:**
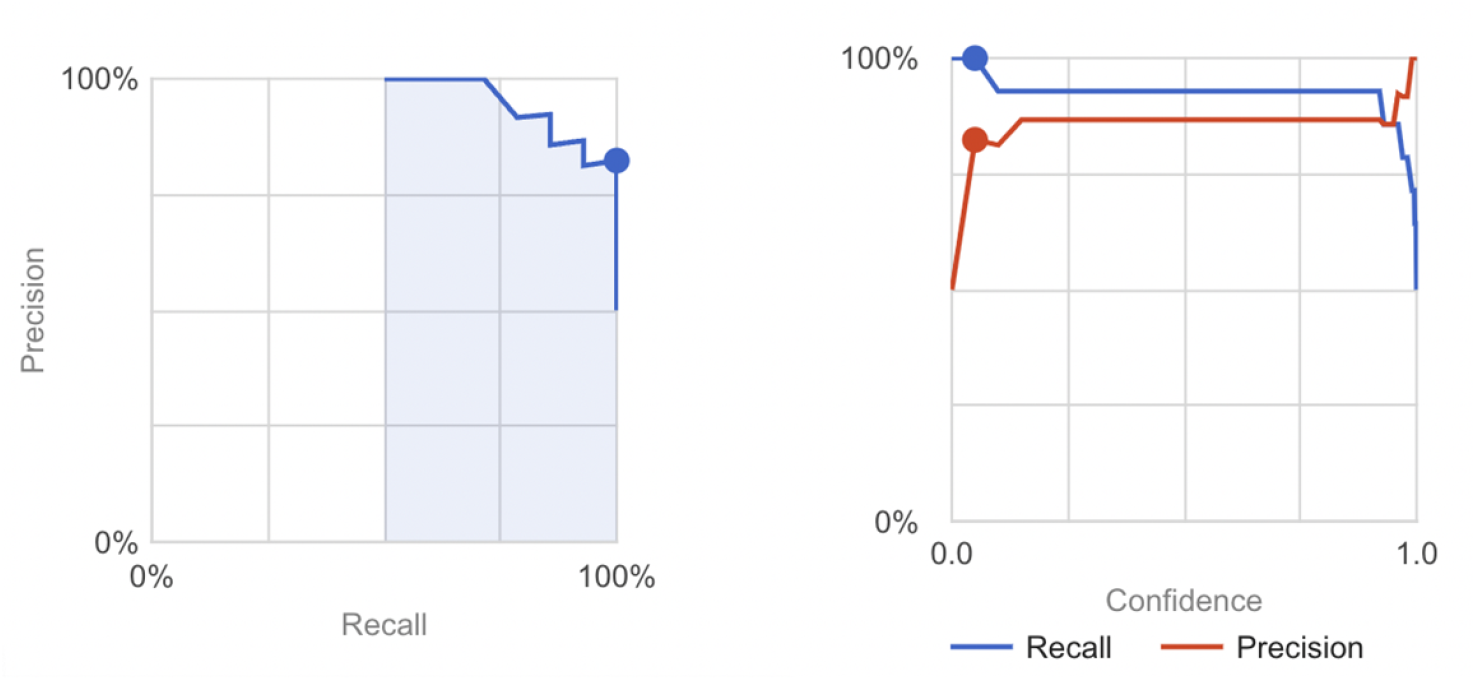
Precision v/s Recall Trade-off for 0.05 confidence threshold.

Though sensitivity remains unchanged in this case, Recall is maximised, albeit compromising the value of precision. Ultimately, Sensitivity and Recall maximisation is the goal of the Clinical Validation, ensuring that every Down Syndrome Positive child is successfully diagnosed and treated. A slight reduction in precision is tolerable since false positives can be ruled out through further confirmatory testing.

## DISCUSSION

This non-experimental, cross-sectional study suggests that such a diagnostic aid can be extremely useful in detecting Down Syndrome in India. Not only does this diagnostic tool ensure universal access to Machine Learning technology for the most disadvantaged sections of Indian society, but it is also extremely easy to use since clicking and uploading a photograph is a common skill today. Since the model provides a preliminary diagnosis instantaneously, timely treatment and recommendation for specialised healthcare is delivered on an accelerated timeline, significantly improving the probability of survival. Rather than universal testing, resources can be deployed to confirm the diagnosis and treat high-risk children first and this tool can be used to screen for high-risk children.

This diagnostic tool’s ideal use case is as a preliminary screening tool in the hands of laymen and healthcare workers in remote areas. On risk assessment, we can ensure timely care by referral to secondary and tertiary care centres. We believe that this approach can reduce the mortality due to Down Syndrome, but to confirm this large well designed population studies will be needed as randomized controlled trials are not feasible for rare diseases.

This study provides valuable insights into Down Syndrome detection for children of Indian (Southeast Asian) origin and how Down Syndrome may be diagnosed on the basis of their unique facial dysmorphic features. The model is better suited to address features and inflections more commonly associated with Indian children and even to assign more relevant weightage to each facial characteristic that may point to Down’s.

India is a country where health equity is a major issue. Even though the affordability is much higher when compared to other countries especially developed nations, there is a huge discrepancy in not only to access to healthcare but healthcare in general. A retrospective observational study conducted in Ireland showed out of 88% clinically suspected patients, only 55% were diagnosed within the first 48 hours after birth, and the number of correctly identifying patients with mosaicism fell to 37.5% (23). Another study from the Netherlands showed that only 76.5% of patients were diagnosed within the first 24 hours after birth (24). These studies also reported early detection in hospital-based deliveries than home deliveries. This data is from countries where the healthcare system is much more equitable, and the checks were done at multiple levels by various healthcare professionals. Study done by Hindley and Medakkar showed that only 68% of all the patients who were suspected to have down syndrome and were referred for further testing, were actually positive (25).

Amongst postnatal tests, Karyotyping is the most frequently prescribed confirmatory test and costs ∼₹2,500-3,000. ‘Interphase Fluorescent in situ Hybridization’ or FISH test provides comparable accuracy at the cost of ∼₹5,000-6,000. In contrast, our App-based fundamental testing will be free for users. Compared to traditional diagnostics, our App has several advantages, especially for the underserved sections of society where it can be used as a screening tool due to the easy availability of mobile phones and the internet, even in the most remote areas. Using this tool does not require any technical skills except for clicking a photograph. This proves to be a very accessible and easy-to-use DIY solution for the masses. Furthermore, this test is non-invasive and painless and can be conducted without posing any risk to the neonate. A preliminary, instantaneous diagnosis is handed out immediately, indicating the risk of having Down Syndrome. This tool also ensures that rather than universal testing, resources can be deployed to confirm the diagnosis of high-risk children, which might lead to optimal use of resources. Karyotyping may now be conducted succinctly, with higher risk patients prioritised.

The Cloud ML Vision Model has been shown to be a reliable tool for early diagnosis of Down Syndrome in neonates and infants of Indian origin, through the identification of facial dysmorphic features. This Indo-specific model provides more relevant and accurate results when compared to other models. The model can be incorporated into a mobile or web application that can be used by parents or as part of standard neonatal screening protocols in maternity centres and even in remote or rural areas. Children identified as high risk by the app can be referred to district or government hospitals for confirmation of diagnosis and treatment. It is important to note that this model should be used as a supplementary tool and not as a replacement for doctors.

## Data Availability

The deindetified data will be available on request.

## REFERENCES

1. Bhide P, Kar A. A national estimate of the birth prevalence of congenital anomalies in India: systematic review and meta-analysis. BMC Pediatr. 2018 May 25;18:175.

2. Asim A, Kumar A, Muthuswamy S, Jain S, Agarwal S. “Down syndrome: an insight of the disease.” J Biomed Sci. 2015 Jun 11;22(1):41.

3. Nahar R, Kotecha U, Puri RD, Pandey RM, Verma IC. Survival Analysis of Down Syndrome Cohort in a Tertiary Health Care Center in India. Indian J Pediatr. 2013 Feb 1;80(2):118–23.

4. Baxi V, Edwards R, Montalto M, Saha S. Digital pathology and artificial intelligence in translational medicine and clinical practice. Mod Pathol. 2022;35(1):23–32.

5. Shah BV, Nimbalkar SM. Artificial intelligence and Neonatology. NNF. 2021;1(2):8–11.

6. Cornejo JYR, Pedrini H, Machado-Lima A, Nunes F de L dos S. Down syndrome detection based on facial features using a geometric descriptor. J Med Imaging. 2017 Oct;4(4):044008.

7. Introduction to built-in algorithms | AI Platform Training [Internet]. Google Cloud. [cited 2023 Jan 20]. Available from: https://cloud.google.com/ai-platform/training/docs/algorithms

8. Kasthuri A. Challenges to Healthcare in India - The Five A’s. Indian J Community Med Off Publ Indian Assoc Prev Soc Med. 2018;43(3):141–3.

9. Sheth V. 5 reasons India’s ‘Missing Middle’ is struggling to access quality healthcare. The Times of India [Internet]. [cited 2023 Jan 19]; Available from: https://timesofindia.indiatimes.com/blogs/voices/5-reasons-indias-missing-middle-is-struggling-to-access-quality-healthcare/?source=app&frmapp=yes

10. India - Rural Population - 2023 Data 2024 Forecast 1960-2021 Historical [Internet]. [cited 2023 Jan 19]. Available from: https://tradingeconomics.com/india/rural-population-percent-of-total-population-wb-data.html

11. Ou CY, Yasmin M, Ussatayeva G, Lee MS, Dalal K. Maternal Delivery at Home: Issues in India. Adv Ther. 2021 Jan;38(1):386–98.

12. de Jonge E, Azad K, Hossen M, Kuddus A, Manandhar DS, van de Poel E, et al. Socioeconomic inequalities in newborn care during facility and home deliveries: a cross sectional analysis of data from demographic surveillance sites in rural Bangladesh, India and Nepal. Int J Equity Health. 2018 Aug 15;17(1):119.

13. Verma A, Cleland J. Is newborn survival influenced by place of delivery? A comparison of home, public sector and private sector deliveries in India. J Biosoc Sci. 2022 Mar;54(2):184–98.

14. Lou S, Nielsen CP, Hvidman L, Petersen OB, Risør MB. Coping with worry while waiting for diagnostic results: a qualitative study of the experiences of pregnant couples following a high-risk prenatal screening result. BMC Pregnancy Childbirth. 2016 Oct 21;16(1):321.

15. Shaw SW, Hsu JJ, Lee CN, Hsiao CH, Chen CP, Hsieh TT, et al. First- and Second-trimester Down Syndrome Screening: Current Strategies and Clinical Guidelines. Taiwan J Obstet Gynecol. 2008 Jun 1;47(2):157–62.

16. Quinlan MP. Amniocentesis: Indications and Risks. AMA J Ethics. 2008 May 1;10(5):304–6.

17. Caughey AB, Kaimal AJ, Odibo AO. Cost-Effectiveness of Down Syndrome Screening Paradigms. Clin Lab Med. 2010 Sep 1;30(3):629–42.

18. Prenatal Testing for Down Syndrome [Internet]. ucsfhealth.org. [cited 2023 Jan 19]. Available from: https://www.ucsfhealth.org/Education/Prenatal Testing for Down Syndrome

19. Fluorescence In Situ Hybridization (FISH) [Internet]. Genome.gov. [cited 2023 Jan 19]. Available from: https://www.genome.gov/genetics-glossary/Fluorescence-In-Situ-Hybridization

20. Precision and recall. In: Wikipedia [Internet]. 2022 [cited 2023 Jan 20]. Available from: https://en.wikipedia.org/w/index.php?title=Precision_and_recall&oldid=1069330375

21. Machine Learning Model Accuracy [Internet]. DataRobot AI Platform. [cited 2023 Jan 20]. Available from: https://www.datarobot.com/wiki/accuracy/

22. What is the Confidence Threshold, and what value should I set for it? [Internet]. Helpshift Knowledge Base. [cited 2023 Jan 19]. Available from: https://support.helpshift.com/kb/article/what-is-the-confidence-threshold-and-what-value-should-i-set-for-it/

23. Devlin L, Morrison PJ. Accuracy of the clinical diagnosis of Down syndrome. Ulster Med J. 2004 May;73(1):4–12.

24. de Groot-van der Mooren MD, Gemke RJBJ, Cornel MC, Weijerman ME. Neonatal diagnosis of Down syndrome in the Netherlands: suspicion and communication with parents. J Intellect Disabil Res. 2014;58(10):953–61.

25. Hindley D, Medakkar S. Diagnosis of Down’s syndrome in neonates. Arch Dis Child - Fetal Neonatal Ed. 2002 Nov 1;87(3):F220–1.

